# Low blood sodium increases risk and severity of COVID-19: a systematic review, meta-analysis and retrospective cohort study

**DOI:** 10.1101/2020.05.18.20102509

**Authors:** Yi Luo, Yirong Li, Jiapei Dai

**Affiliations:** Wuhan Institute for Neuroscience and Neuroengineering, South-Central University for Nationalities, Wuhan 430074, China; Department of Laboratory Medicine, Zhongnan Hospital of Wuhan University, Donghu Road 169, Wuhan 430071, China; College of Life Sciences, South-Central University for Nationalities, Wuhan 430074, China

## Abstract

**Background:** **Novel coronavirus (SARS-CoV-2) infects human lung tissue cells through angiotensin-converting enzyme-2 (ACE2), and the body sodium is an important factor for regulating the expression of ACE2. Through a systematic review, meta-analysis and retrospective cohort study, we found that the low blood sodium population may significantly increase the risk and severity of SARS-CoV-2 infection**.

**Methods:** **We extracted the data of serum sodium concentrations of patients with COVID-19 on admission from the articles published between Jan 1 and April 28, 2020, and analyzed the relationship between the serum sodium concentrations and the illness severity of patients. Then we used a cohort of 244 patients with COVID-19 for a retrospective analysis**.

**Results:** **We identified 36 studies, one of which comprised 2736 patients.The mean serum sodium concentration in patients with COVID-19 was 138.6 mmol/L, which was much lower than the median level in population (142.0). The mean serum sodium concentration in severe/critical patients (137.0) was significantly lower than those in mild and moderate patients (140.8 and 138.7, respectively). Such findings were confirmed in a retrospective cohort study, of which the mean serum sodium concentration in all patients was 137.5 mmol/L, and the significant differences were found between the mild (139.2) and moderate (137.2) patients, and the mild and severe/critical (136.6) patients. Interestingly, such changes were not obvious in the serum chlorine and potassium concentrations**.

**Conclusions:** **The low sodium state of patients with COVID-19 may not be the consequence of virus infection, but could be a physiological state possibly caused by living habits such as low salt diet and during aging process, which may result in ACE2 overexpression, and increase the risk and severity of COVID-19. These findings may provide a new idea for the prevention and treatment of COVID-19**.

## Introduction

A global pandemic of the novel coronavirus (2019-nCoV) –infected pneumonia (NCIP) or coronavirus disease 2019 (COVID-19) has caused great impact on the world economy and human health and life. Clinical research findings have demonstrated that the infection and transmission of this virus has a wide range of susceptibility in populations, but also highlights some of the following characteristic:^1–7^the elderly and people with basic diseases and underlying conditions such as cardiovascular diseases, diabetes, chronic kidney diseases and obesity, etc., are prone to develop to be severe/critical situations after infection; the young and middle-aged people after infection usually have mild symptoms and less chance of forwarding to the severe/critical conditions; the minors are less likely to be infected and onset; there are asymptomatic virus carriers; the mortality rate is related to countries, regions and races.^8^ Up to now, the exact reasons and mechanisms that cause such epidemic characteristic have not been clear, and no specific therapeutic drugs and reliable vaccines have been developed and used. Therefore, it is very important to find the key susceptible factors for the prevention and control of this deadly epidemic disease.

The gene sequence of SARS-CoV-2 is highly homologous with that of SARS-CoV.^9, 10^ Previous studies have found that such types of viruses mainly enter host cells through the receptor angiotensin-converting enzyme 2 (ACE2),^11, 12^and then replicates and releases new viruses from the host cells, resulting in virus overload in the body, and finally leading to different degree of damage to tissues and organs, especially in lung, through a series of complex pathophysiological mechanisms.

A large number of epidemiological studies have demonstrated that low salt diet plays a role in the prevention and reduction of cardiovascular and cerebrovascular diseases.^13–19^ Therefore; it is widely promoted as a healthy diet habit in developed countries and some developing countries. In animal studies, it was found that the decrease of salt intake can upregulate the expression of ACE2 through the activation of the renin-angiotensin-aldosterone system (RAAS).^20, 21^ Therefore, we could speculate that the long-term low salt diet or the condition of sodium insufficiency may result in the low body sodium and the overexpression of ACE2 in lower respiratory tract cells, which could increase the susceptibility and pathogenicity of SARS-CoV-2 infection. In this study, we aimed to find a key risk factor for SARS-CoV-2 epidemic by investigating the relationship between the blood sodium concentration and the severity of patients with COVID-19 through a systematic reviews, meta-analysis and retrospective cohort study.

## Methods

### Systematic review and meta-analysis

#### Search strategy and selection criteria

We did a systematic review and meta-analysis from the observational and retrospective cohort studies to compare the serum sodium concentrations of confirmed patients with COVID-19 in relation to the severity of disease according to PRISMA guidelines.^22^ MedRxiv and BioRxiv databases, PubMed and the Web of Science Core Collection (Clarivate Analytics) were used for searching articles published between Jan 1 and April 28, 2020 using the keyword “sodium” in combination with other keywords “SARS-CoV-2”, “COVID-19”, “Coronavirus”, “nCoV”, “HCoV” or “NCIP”. Detailed search strategies were presented in Figure 1. After removing duplicates, two researchers were assigned to independently screen the titles and abstracts, and then examine the full text. Any questions with conflict were resolved by discussion. Inclusion criteria were as follows: (1) any study must contain the information about serum biochemical test of electrolytes on admission, (2) restriction language to English only, and (3) studies that presented the severity of disease in relation to the serum sodium concentrations or allowed us to evaluate the severity of disease by using the clinical information, experimental tests and imaging data, etc., presented in the reports according to the criteria as mentioned below in the retrospective cohort study. Exclusion criteria were (1) data that could not be reliably extracted, (2) reviews, editorials, comments, expert opinions, case reports or articles with small number of cases (≤ 10).

**Figure 1.**
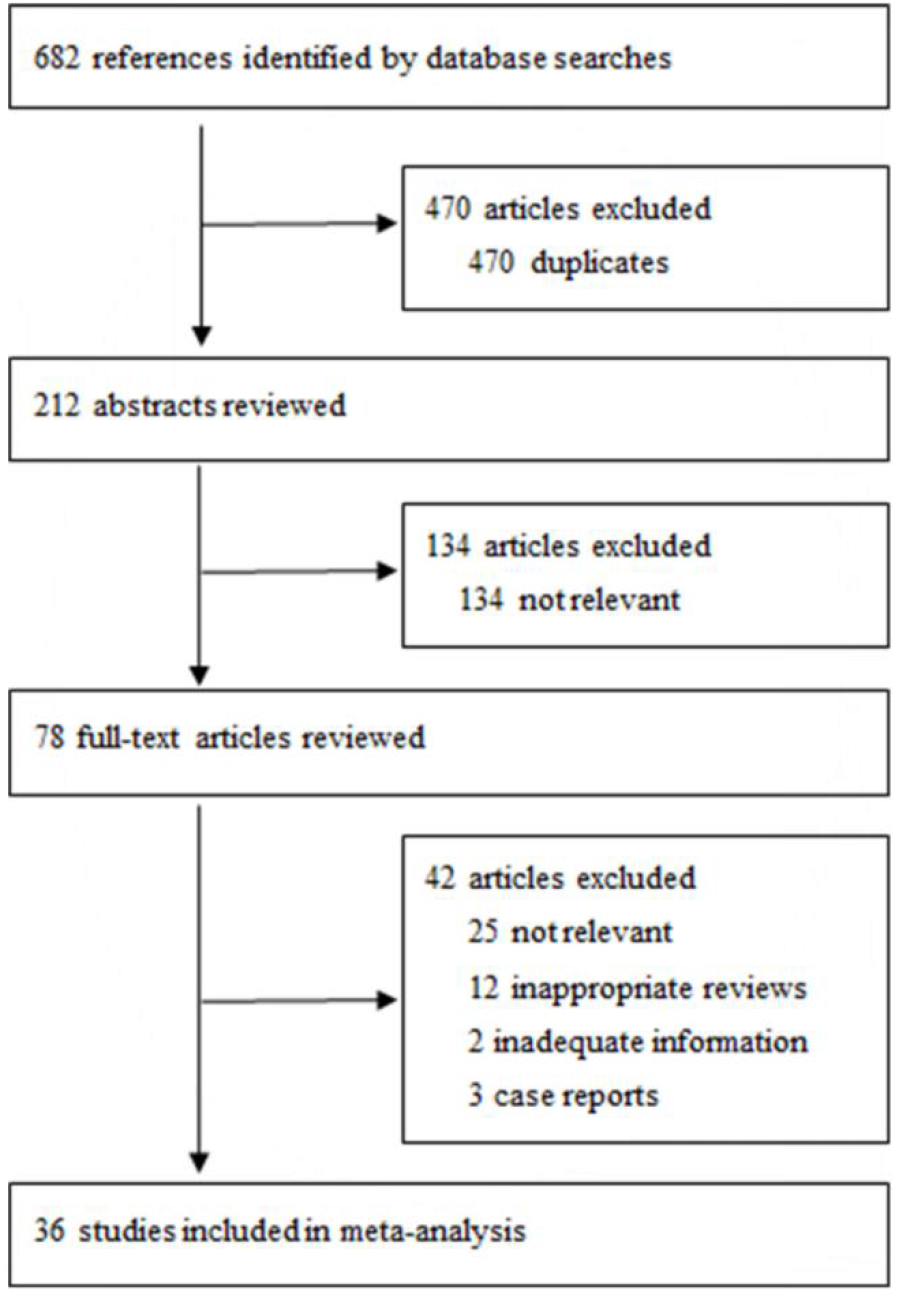
Study selection in systematic review and meta-analysis. Serum sodium in patients with COVID-19.

### Retrospective cohort study

#### Study design and participants

The retrospective cohort study included a cohort of 244 COVID-19 inpatients aged from 7 to 96 years old and 59 non-COVID-19 inpatients (control) aged from 10–90 years old from Zhongnan Hospital of Wuhan University (Wuhan, China). All patients who were diagnosed with COVID-19 according to WHO interim guidance were screened from Dec 31, 2019 to Feb 27, 2020 (control patients from Dec 1, 2019 to Jan 25, 2020). This case series was approved by the Zhongnan Hospital of Wuhan University for Ethics Committee (No. 2019125).

## Definition of severity

The severity of patients with COVID-19 was defined at admission based on the criteria established by China’s National Health Commission.^23^ Mild (I): Minor symptoms only, without evidence for pneumonia by chest X-ray; Moderate (II): Fever and respiratory symptoms are present, and there is evidence for pneumonia by chest X-ray; Severe (III): Defined by any of the following conditions: (1) Dyspnoea, respiratory rate 30 /min, (2) resting hypoxia SaO2 ≤93%, and (3) PaO2/FiO2 ≤300 mmHg; Critical (IV): The presence of any of the following conditions: (1) Respiratory failure, require mechanical ventilation, (2) shock, and (3) other acute organ failure.

## Laboratory procedures

Methods for laboratory confirmation of COVID-19 infection have been described elsewhere.^6^ COVID-19 detection was done in respiratory specimens by next-generation sequencing or real-time RT-PCR methods. Serum biochemical test of electrolytes at admission was carried out with other tests including renal and liver function, creatine kinase, lactate dehydrogenase, myocardial enzymes, interleukin-6 (IL-6), serum ferritin, and procalcitonin, etc.

## Data collection

For the systematic review and meta-analysis, the data for laboratory test of serum sodium, chloride and potassium concentrations, and the clinical severity of patients on admission were collected, and the concentration is represented as median (IQR, interquartile range) or mean (SD, standard deviation) depending on the reports. For the retrospective cohort study, epidemiological, demographic, clinical, laboratory, treatment, and outcome data were extracted from electronic medical records, and only basic information such as age, gender, clinical severity of patients and serum sodium, potassium and chloride concentrations on admission were selected for data analysis. All data were checked independently by two researchers.

## Data analysis

For the systematic review and meta-analysis, median or mean values of serum sodium, chloride and potassium concentrations from each report were considered as an independent variable for statistical analysis, and an unpaired *t*-test was used to compare the differences between the groups related to the severity of disease. For the retrospective cohort study, one-way ANOV, followed by Tukey’s multiple comparisons test, was used to compare the differences among groups or an unpaired t-test was used if necessary. To assess the association between sodium, chloride or potassium concentration, and the severity of patients with COVID-19, Spearmann’s rank correlation coefficient was used. P<0.05 was defined as showing statistical significance of differences. All analyses were performed by using GraphPad Prism 7 software.

## Results

For the systematic review and meta-analysis, we identified 682 papers published between Jan 1 and April 28, 2020. After removing unsuitable studies (duplicates, not relevant, inappropriate reviews, inadequate information and case reports), 36 studies had adequate data that were represented in the dataset (Table S1) for further analysis. These studies included 32 observational (or cohort) reports from different regions in China, 3 from the United States (US), and 1 from Iran. The smallest sample size included 12 patients and the largest one comprised 2736 patients. 15 of 36 studies (44.4%) provided the normal range of serum sodium concentration, which mostly were 137–147 mmol/L (Table S1).

We found that the serum sodium concentrations in patients with COVID-19 on admission presented the characteristic as follows (Table 1, Figure 2): regardless of the disease severity, the mean serum sodium concentration was 138.6 mmol/L (138.6±2.5, n=31), which was significantly lower than the median level (142.0) of normal reference range (137.0–147.0); the mean serum sodium concentration in severe/critical patients was 137.0 mmol/L (137.0±2.02, n=25), close to the lower level of normal range, and was significantly lower than those in mild (140.8±4.0, n=7) and moderate (138.7±2.35, n=23) patients (*P* = 0.0019 and *P*=0.0141, respectively), but there was no significant difference between the mild and moderate patients (*P*=0.0927) (Table 1, Figure 2A); some studies showed that the serum sodium concentrations in patients who died after admission were even lower than that in recovered patients (Table S1, refs 30); the serum sodium concentrations in severe/critical patients with tuberculosis (Table S1, ref 16) were very low (131.67±3.08); Three studies from the United States demonstrated that the median level of serum sodium concentrations in patients with COVID-19 was approximately 136.0 mmol/L (135.5–137.0), close to the lower level of normal range (Table S1, refs 23, 31 and 35); The median serum sodium concentration from 34 paediatric patients aged from 1 to 144 months (median 33) with COVID-19 was 139.0 mmol/L (137.88–141.13), which is lower than the median level of normal range (Table S1, ref 17). The serum sodium concentrations were significantly correlated to the illness severity of patients (Table 1, Figure 2B, r=–0.4558, p=0.0005).

**Table 1.** The results of systematic review and meta-analysis.

| Table 1 The results of systematic review and meta-analysis |  |  |  |  |  |
| --- | --- | --- | --- | --- | --- |
|  | Severity | Mean ± SD | Number of | Comparison | P value |
| Sodium (Na) | All | 138.6±2.5 | 31 | All vs. Mild | 0.0774 |
|  | Mild | 140.8±4.0 | 7 | All vs. Moderate | 0.9531 |
|  | Moderate | 138.7±2.35 | 23 | All vs. Severe/Critical | 0.0139 |
|  | Severe/Critical | 137.0±2.02 | 25 | Mild vs. Moderate | 0.0927 |
|  | Correlation | r= -0.4558 |  | Na concentration vs Severity | 0.0005 |
| Chlorine (Cl) | All | 101.8±2.25 | 10 | All vs. Moderate | 0.7769 |
|  | Mild | 102.9-103.4 | 2 | All vs. Severe/Critical | 0.5839 |
|  | Moderate | 102.1±2.39 | 9 | Moderate vs. Severe/Critical | 0.419 |
|  | Severe/Critical | 101.3±2.06 | 10 |  |  |
|  | Correlation | r= -0.2817 |  | Cl concentration vs Severity | 0.216 |
| Potassium (K) | All | 4.01±0.20 | 28 | All vs. Mild | 0.7673 |
|  | Mild | 3.99±0.18 | 7 | All vs. Moderate | 0.5866 |
|  | Moderate | 3.99±0.19 | 21 | All vs. Severe/Critical | 0.1199 |
|  | Severe/Critical | 3.90±0.28 | 23 | Mild vs. Moderate | 0.942 |
|  | Correlation | r= -0.1523 |  | K concentration vs Severity | 0.2861 |
| SD: Standard deviation |  |  |  |  |  |

**Figure 2.**
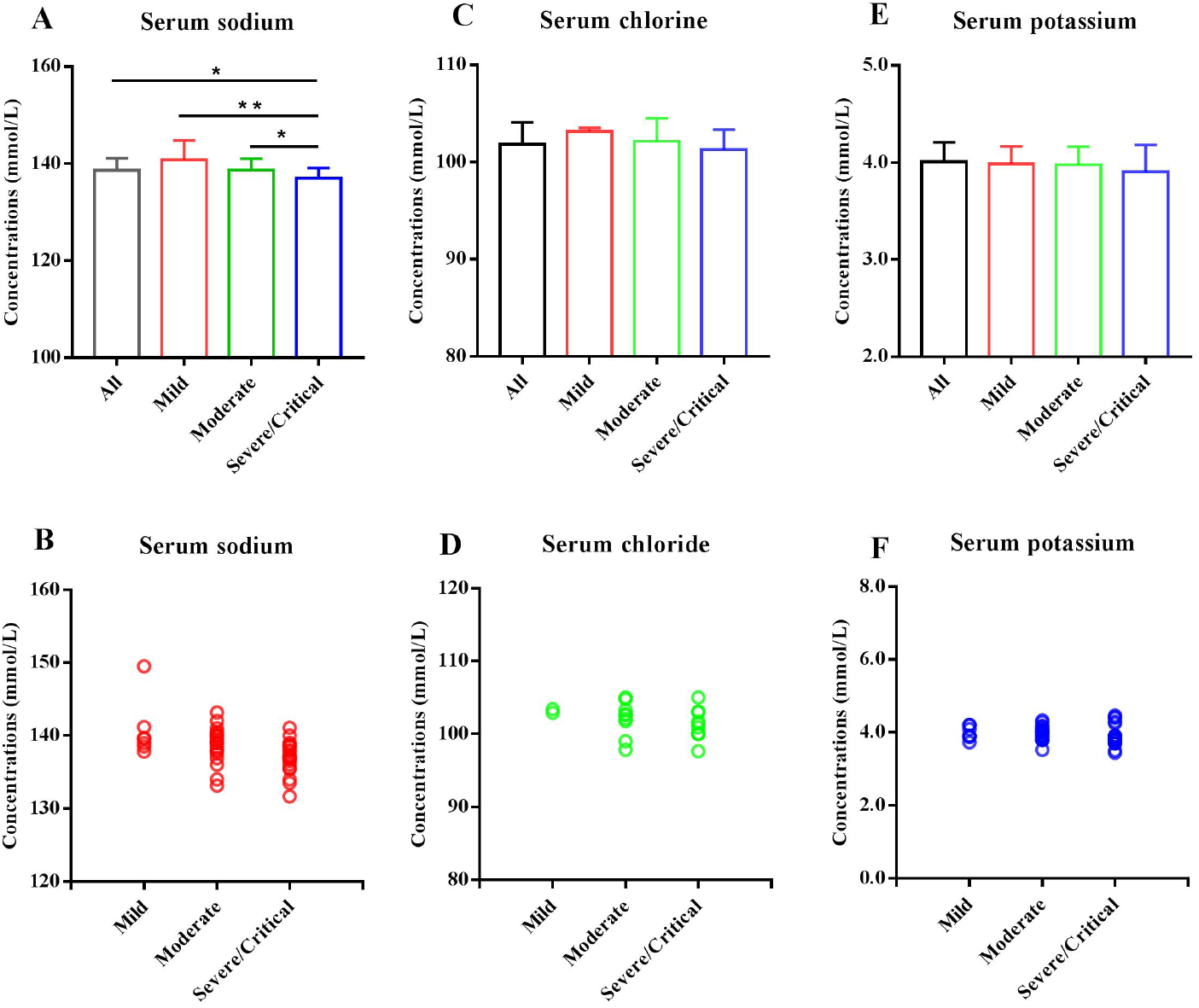
Comparisons in serum sodium, chloride and potassium concentrations related to the severity of patients with COVID-19 based on the data of systematic review and meta-analysis. The mean serum sodium concentrations in all (n=31), mild (n=7), moderate (n=23) and severe/critical (n=25) patients with COVID-19, and the significant differences were found between the groups as indicated by bar lines and asterisks (A). The concentrations of serum sodium were significantly correlated to the illness severity of patients (B). The mean serum chloride concentrations in all (n=10), mild (n=2), moderate (n=9) and severe/critical (n=10) patients with COVID-19 (C). The concentrations of serum chloride were not correlated to the illness severity of patients (D). The mean serum potassium concentrations in all (n=28), mild (n=7), moderate (n=21) and severe/critical (n=23) patients with COVID-19 (E). There were no differences in serum chloride and potassium concentrations among the different groups. The serum potassium concentrations were also not correlated to the illness severity of patients (F). Data are shown as the mean ± SD. n=the number of studies. Asterisks indicate a significant difference in different groups. \**P*<0.05, \*\**P*<0.01.

In addition, we analyzed the serum chloride and potassium concentrations related to the disease severity. The mean serum chlorine concentrations in all, mild, moderate and severe/critical patients were 101.8 mmol/L (101.8±2.25, n=10), 103.15 mmol/L (102.9 and 103.4, respectively, n=2), 102.1 mmol/L (102.1±2.39, n=9) and 101.3 mmol/L (101.3±2.06, n=10), respectively, and no statistical differences were found between the comparable groups (Table 1, Figure 2C), while the mean serum potassium concentrations in all, mild, moderate and severe/critical patients were 4.01 mmol/L (4.01±0.20, n=28), 3.99 mmol/L (3.99±0.18, n=7), 3.99 mmol/L (3.99±0.19, n=21) and 3.90 mmol/L (3.90±0.28, n=23), respectively, and there were no statistical differences between the groups (Table 1, Figure 2E). The serum chloride and potassium concentrations were not correlated to the severity of patients (Table 1, Figure 2D, F).

For the retrospective cohort analysis, we collected data of 244 laboratory-confirmed COVID-19 patients and 59 non-Covid-19 patients as control from a single center (Zhongnan Hospital of Wuhan University) between Dec 30, 2019 and Feb 27, 2020 (from Dec 1, 2019 to Jan 9, 2020 for control). The data of age, gender and severity of patients with and without COVID-19 were summarized in Table 2. The average age was 57.49 years (57.49±19.13) from 7–96 years in the patients with COVID-19, and 51.07 years (51.07±19.21) from 10–90 years in the control group. 127 of 244 (52.05%) were male, 117 (47.95 %) were female, and 7 were minors (2.87 %) in the COVID-19 group. The numbers of mild, moderate, severe and critical patients were 65, 61, 77, and 41, respectively. We have obtained very similar results as those in the systematic review and meta-analysis (Table 2, Figure 3): the mean serum sodium concentration in all patients was 137.5 mmol/L (137.5±4.12, n=244, Table 2, Figure 3A, D), which was much lower than the median value of normal reference range (142.0 mmol/L, 137.0 –147.0, Table 2) and significant lower than the mean level of control patients (138.9±2.77, n=59, *p*=0.0443, Table 2, Figure 3D); the mean serum sodium concentration in 60 years and older patients was 136.6 mmol/L (136.6±4.62, n=120), which was significant lower than that in patients under 60 years (138.3±3.411, n=124, *P*=0.0019, Table 2, Figure 3B); there was significant difference between male and female in the mean serum sodium concentrations (136.7±4.25 and 138.3±3.82, *P*=0.0021, Table 2, Figure 3C), however, there were no difference in gender and age (<60 years and ≥60 years) in control patients (Table 2); the mean serum sodium concentrations in the mild, moderate and severe/critical patients were 139.2 mmol/L (139.2±2.72, n=65), 137.2 mmol/L (137.2±3.72, n=61), and 136.6 mmol/L (136.6±4.66, n=118), respectively, and the differences between the mild and moderate groups as well as the mild and severe/critical groups were significant (*p*=0.0330 and *p*=0.0003, respectively), but there was no significant difference between the moderate and severe/ critical groups (*p*=0.7886) (Table 2, Figure 3D); The serum sodium concentrations were significantly correlated to the illness severity of patients (Table 2, Figure 3E, r=–0.2462, *p*=0.0001).

**Table 2.**
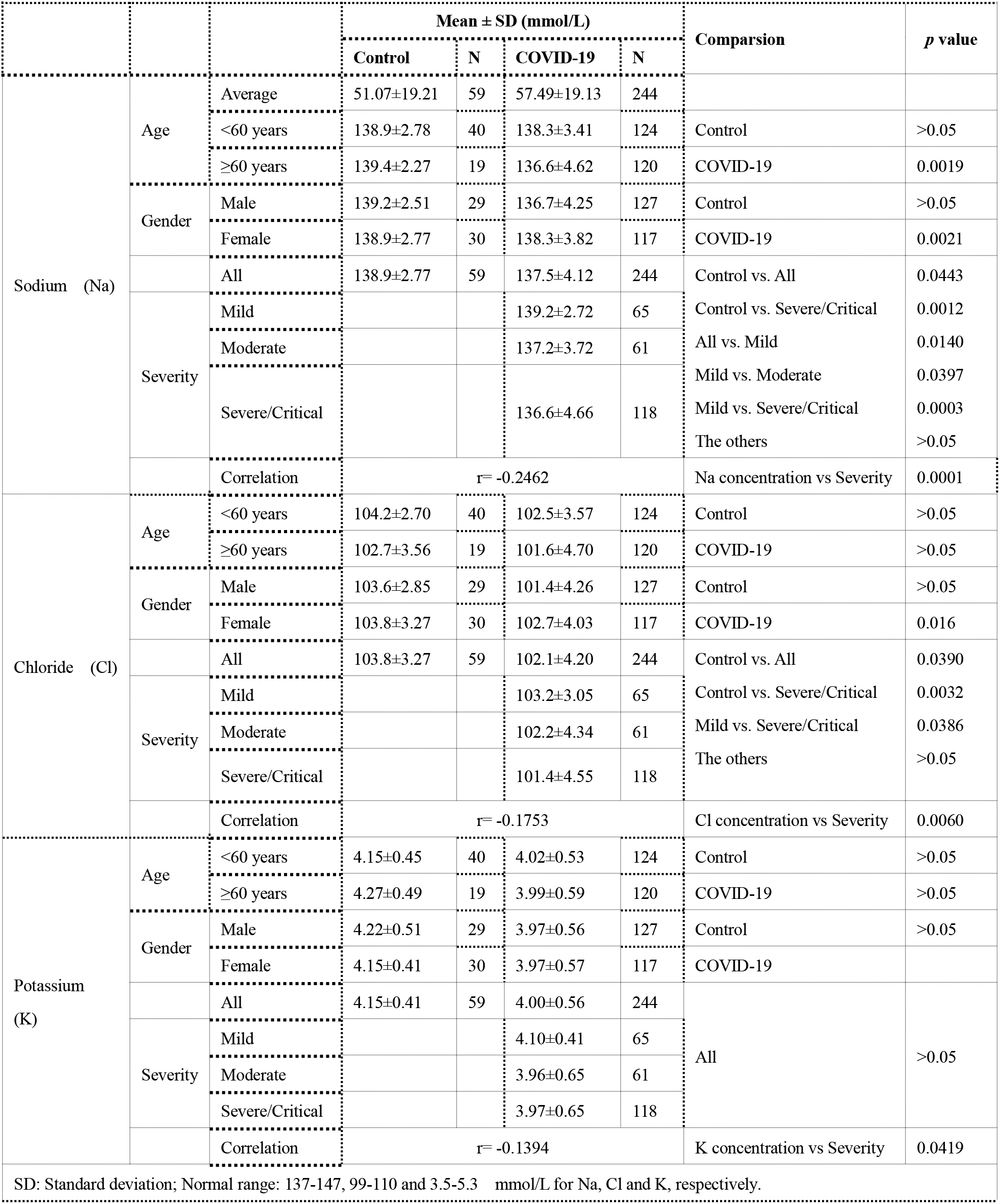
The results of retrospective cohort study.

**Figure 3.**
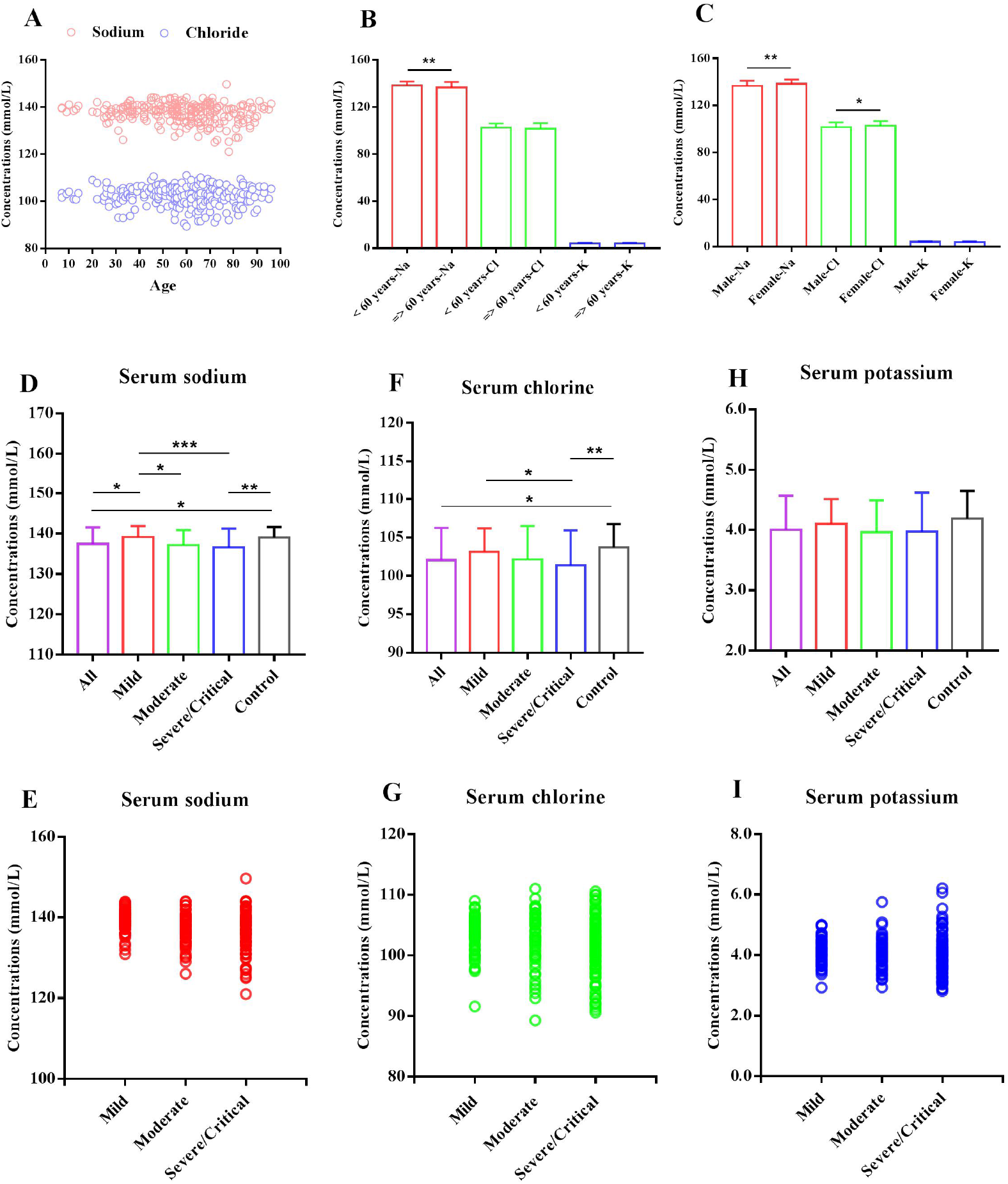
Comparisons in serum sodium, chloride and potassium concentrations related to the severity of patients with COVID-19 based on the data of retrospective cohort study. The age-dependent distribution of serum sodium and chloride concentrations in 244 patients with COVID-19 (A). The serum sodium, chloride and potassium concentrations in patients with COVID-19 related to age (<60 vs ≥60 years). A significant difference was found in the serum sodium concentrations, but not in serum chloride and potassium concentrations (B). The serum sodium, chloride and potassium concentrations in patients with COVID-19 related to gender. The significant differences were found in the serum sodium and chloride concentrations, but not in serum potassium concentrations (C). The mean serum sodium (D), chloride (F) and potassium (H) concentrations in control patients (n=49), and all (n=244), mild (n=65), moderate (n=61) and severe/critical (n=118) patients with COVID-19. The significant differences were found between the groups as indicated by bar lines and asterisks. The correlations between the serum sodium, chloride and potassium concentrations and the illness severity of patients with COVID-19 were shown in F, G and I, respectively. The concentrations of serum sodium and chloride, but not serum potassium were significantly correlated to the illness severity of patients. Data are shown as the mean ± SD. n=the number of patients. \**P*<0.05, \*\**P*<0.01, ***P<0.001.

We also analyzed the serum chloride and potassium concentrations in all patients and there were no significant differences in gender (male vs female) and age (≥ 60 vs <60 years) in the non-COVID patients (Table 2). The mean serum chlorine concentrations in all, mild, moderate and severe/critical patients were 102.1 mmol/L (102.1±4.20, n=244), 103.2 mmol/L (103.2±3.05, n=65), 102.2 mmol/L (102.2±4.34, n=61), and 101.4 mmol/L (101.4±4.55, n=118), respectively. There were no significant differences between the groups with the exception of the mild and severe/critical groups (*p*=0.0306) (Table 2, Figure 3F). The serum chloride concentrations were significantly correlated to the severity of patients (Table 2, Figure 3G, r=–0.1753, *p*<0.0060). The mean serum potassium concentrations in all, mild, moderate and severe/critical patients were 4.00 mmol/L (4.00±0.56, n=244), 4.10 mmol/L (4.10±0.41, n=65), 3.96 mmol/L (3.96±0.65, n=61) and 3.97 mmol/L (3.97±0.65, n=118), respectively, and there were no statistical differences between the groups (Table 2, Figure 3H). The serum potassium concentrations were slightly correlated to the severity of patients (Table 2, Figure 3I, r=–0.1394, *p*=0.0419). There was no differences in serum potassium and chlorine concentrations between the 60 years and older patients (3.99±0.59 and 101.6±4.7 mmol/L, respectively, n=120) and the patients under 60 years old (4.02±0.53 and 102.5±3.57 mmol/L, respectively, n=124) (Table 2, Figure 3B).

## Discussion

In this study, we found that the patients infected by SARS-CoV-2 on admission have presented the low blood sodium levels (hyponatremia) that were related to the disease severity. The occurrence of such a condition may not be the consequence of virus infection, but should be a physiological state being existed in the body before virus infection. There were no clear reasons that could cause such a significant reduction of blood sodium concentration because diarrhea and vomiting occurred only in a small number of patients. The fever, a main symptom of patients with CIVID-19, may lead to dehydration and then should increase, but not reduce the blood sodium concentration. Therefore, we could boldly speculate that the low blood sodium population, especially the elderly and people with underlying diseases, may increase the risk and illness severity to SARS-CoV-2 infection, and one possible explanation is that the people companied with long-term hyponatremia may increase the expression of ACE2 in tissue cells, especially in lung cells, through the RAAS.^20, 21^

Hyponatremia is closely related to the incidence and severity of community-acquired pneumonia and perforated acute appendicitis in children.^24–26^ Clinical studies have showed that the elderly are more susceptible to SARS-CoV-2 infection and become more severe than the young and middle aged people, which may be due to the lower blood sodium levels in the elderly population. One of the reasons may be the decrease of the regulation mechanism of sodium ion in the elderly, including the decrease of renal reabsorption function for sodium ion during aging process; however, another key reason may be the result of long-term low sodium diet. For a long time, especially in the developed countries, the strategy of low sodium diet being actively implemented by physicians, medical organizations and public health agencies has played an important role in preventing and controlling hypertension and related diseases,^27,28^ however, some recent evidence suggests that very low sodium intake may actually have adverse effects on human health, which may also preset a very unfavorable state in certain population against SARS-CoV-2 infection. In this study, we found that some young severe patients with CODIV-19 had serious hyponatremia, reinforcing our interpretations. In addition, the low susceptibility of SARS-CoV-2 infection to the minors suggests that the relative high levels of blood sodium may play a role in fighting against the virus infection. Whether an age-dependent change in blood sodium levels is related to increasing expression of ACE2 in human tissue cells, in particular in alveolar cells, remains to be experimentally confirmed.

The presence of high concentration of extracellular sodium ions in lower aquatic organisms, even reaching to 440 mmol/L in squid, may be a natural means for them to fight against various microbial infections besides the realization of physiological functions such as nerve action potential. The blood sodium concentration with an average level of 146.0 mmol/L in bats that can coexist with multiple coronaviruses in vivo is indeed much higher than that in human beings.^29^ In addition, it was found that

North American bat populations, if its blood sodium level was significantly reduced during hibernation, would increase their infection to fungal and develop white nose syndrome causing widespread death of bats.^30^ Such a finding may provide a physiological mechanism for an explanation of coexisting with various coronaviruses in bats, and also indicate a possibility that many asymptomatic SARS-CoV-2 carriers may hold higher blood sodium levels and have lower expression of ACE2 in lower respiratory tract cells.

Three studies from the United States have showed that the median levels of serum sodium in COVID-19 patients were approximately 136.0 mmol/L, which was much lower than an average level of approximately 138.0 mmol/L as reported in most studies from China. It remains to be clarified whether such a difference could provide an explanation for the higher mortality rate after SARS-CoV-2 infection in US as compared to that in China.

There exist two limitations in this study. The data from the systematic review and meta-analysis were mainly extracted from the observational studies in a preprint form due to an unexpected, fast and short-period SARS-CoV-2 pandemic, which would not allow the research works to be published immediately through a traditional review way. However, it could be recognized that the standardization and automatic detection methods for serum electrolytes used in the modern hospitals could ensure the reliability of extracted data from the identified studies. In addition, for the retrospective cohort analysis, although a small cohort of 59 non-COVID-19 inpatients as control have provided very useful control data, however, it would be benefit to our conclusion if a normal control population matched with age and sex, etc.; during the SARS-CoV-2 epidemic period.

In conclusion, we found that people with low serum sodium (hyponatremia) may be related to the susceptibility of SARS-CoV-2 infection and the development of severity of disease. This finding may provide an important idea to prevent the widespread prevalence of this virus or even other types of coronavirus, and to treat the patients. In the epidemic stage, it may be of great significance to properly provide enough sodium intake or maintain blood sodium at a reasonable level for the susceptible population in order to reduce the virus infection, and treat patients through therapeutic sodium supplementation (such as infusion) to prevent from the development of severe condition of disease.

## Contributors

JD had the idea for and designed the study. JD collected the data for systematic review and meta-analysis. YL (first author) and YL collected the data for retrospective cohort study. JD and both of YL had full access to all of the data in the study and take responsibility for the integrity of the data and the accuracy of the data analysis. JD drafted the paper, and JD and both of YL agree to be accountable for all aspects of the work in ensuring that questions related to the accuracy or integrity of any part of the work are appropriately investigated and resolved.

## Data Availability

All data analysed during this study are included in this submitted manuscript (and its supplementary information file). All original documents of clinical data of patients not publicly available but are available from the corresponding author on reasonable request.

## Acknowledgments

This work was partially supported by the research funds of South-Central University for Nationalities (XTZ15014 and CZP 18008).

## Supplementary materials

**Table S1:**
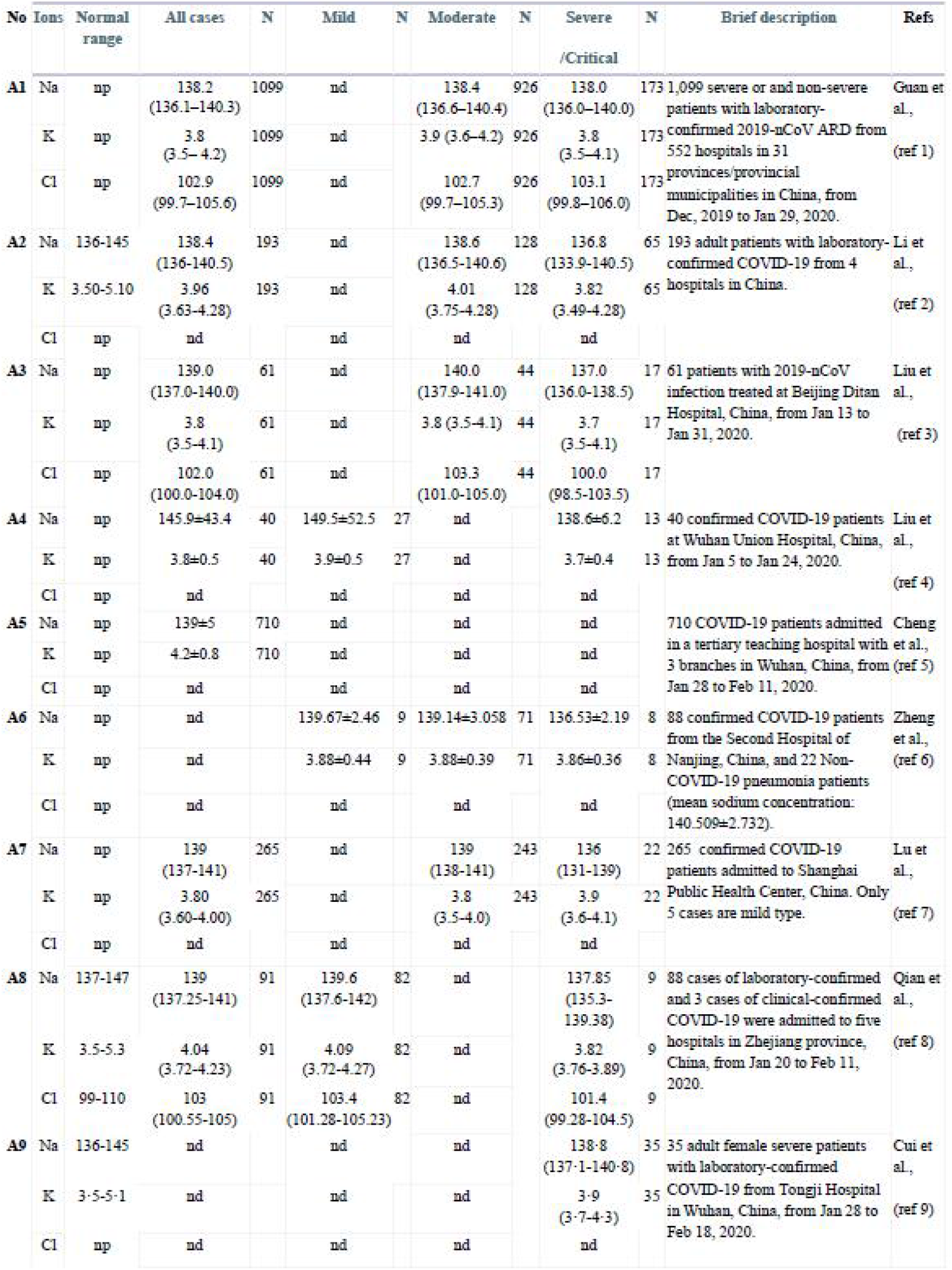
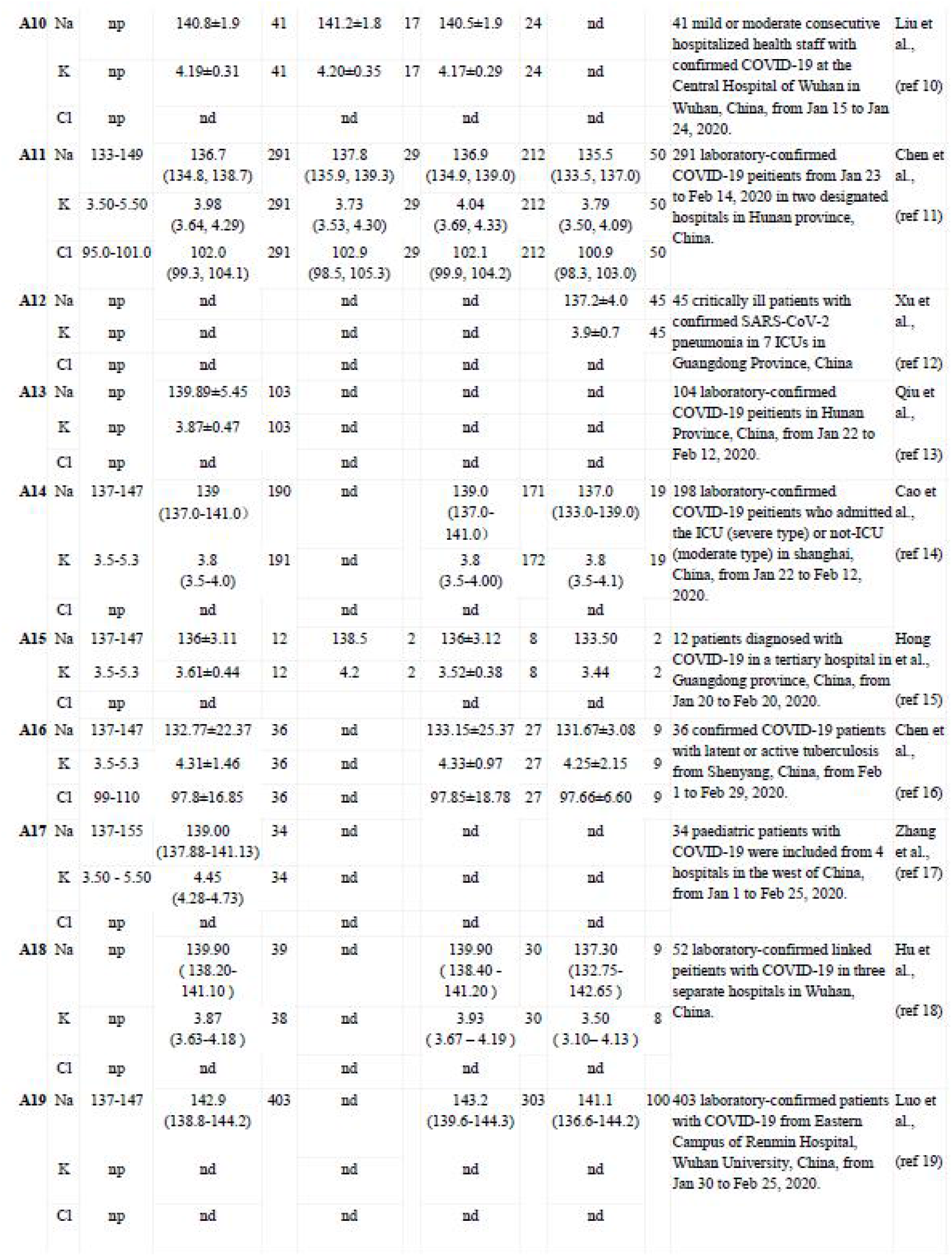
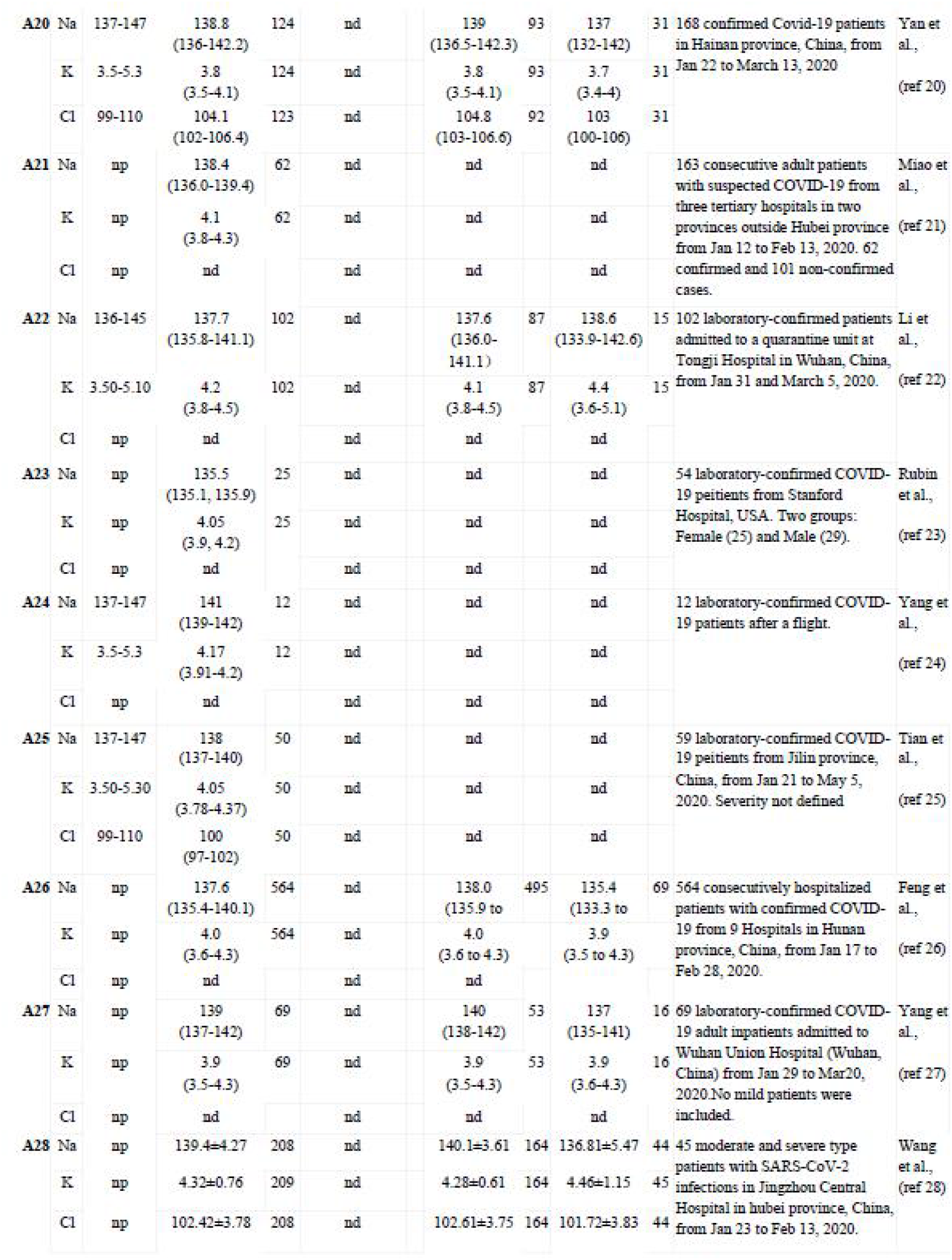
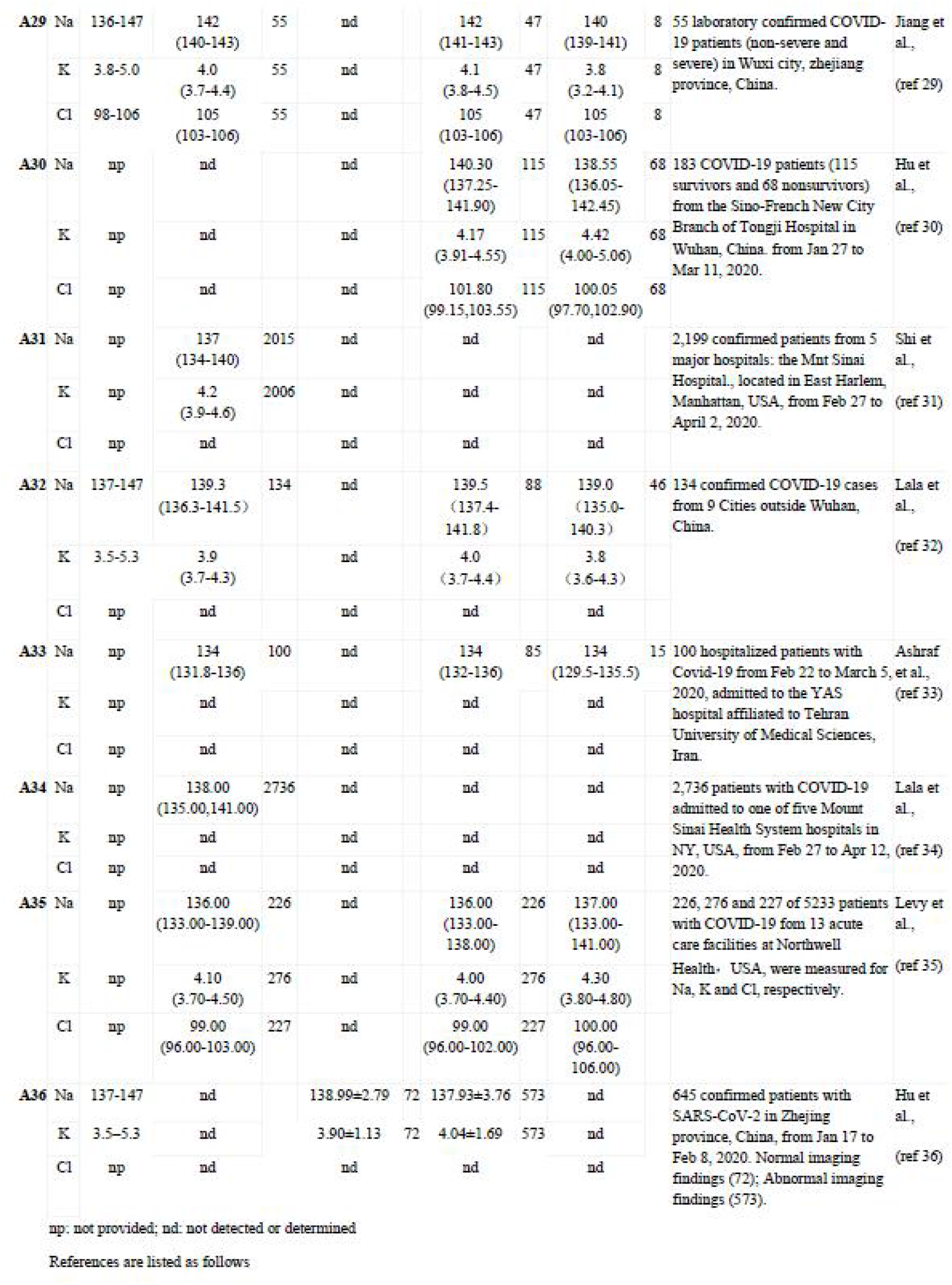
The data of systematics review and meta-analysis

## References

1. Chen N, Zhou M, Dong X, et al. Epidemiological and clinical characteristics of 99 cases of 2019 novel coronavirus pneumonia in Wuhan, China: a descriptive study. Lancet 2020; 395: 507–13.

2. Huang C, Wang Y, Li X, et al. Clinical features of patients infected with 2019 novel coronavirus in Wuhan, China. Lancet 2020; 395: 497–06.

3. Zhu N, Zhang D, Wang W, et al. A novel coronavirus from patients with pneumonia in China, 2019. N Engl J Med 2020; 382: 727–33.

4. Li Q, Guan X, Wu P, et al. Early transmission dynamics in Wuhan, China, of novel coronavirus-infected pneumonia. N Engl J Med 2020; 382: 1199–07.

5. Guan WJ, Ni ZY, Hu Y, et al. Clinical characteristics of coronavirus disease 2019 in China. N Engl J Med 2020; 382:1708–20.

6. Wang D, Hu B, Hu C, et al. Clinical characteristics of 138 hospitalized patients with 2019 novel coronavirus-infected pneumonia in Wuhan, China. JAMA 2020; 323: 1061–69.

7. Zhou F, Yu T, Du R, et al. Clinical course and risk factors for mortality of adult inpatients with covid-19 in Wuhan, china: A retrospective cohort study. Lancet 2020; 395: 1054–62.

8. Wadhera RK, Wadhera P, Gaba P, et al. Variation in COVID-19 hospitalizations and deaths across New York City boroughs. JAMA 2020; 29:e207197.

9. Zhou P, Yang XL, Wang XG, et al. A pneumonia outbreak associated with a new coronavirus of probable bat origin. Nature 2020; 579: 270–73.

10. Lu R, Zhao X, Li J, et al. Genomic characterisation and epidemiology of 2019 novel coronavirus: implications for virus origins and receptor binding. Lancet 2020; 395: 565–74.

11. Ge XY, Li JL, Yang XL, et al. Isolation and characterization of a bat SARS-like coronavirus that uses the ACE2 receptor. Nature 2013; 503: 535–538.

12. Li W, Moore MJ, Vasilieva N, et al. Angiotensin-converting enzyme 2 is a functional receptor for the SARS coronavirus. Nature 2003; 426: 450–454.

13. Ellison RC, Capper AL, Stephenson WP, et al. Effects on blood pressure of a decrease in sodium use in institutional food preparation: The Exeter-Andover Project. Journal of Clinical Epidemiology 1989; 42: 201–08.

14. He FJ, MacGregor GA. Importance of salt in determining blood pressure in children: meta-analysis of controlled trials. Hypertension 2006; 485: 861–69.

15. Strazzullo P, D’Elia L, Kandala NB, Cappuccio FP. Salt intake, stroke, and cardiovascular disease: meta-analysis of prospective studies. BMJ 2009; 339: b4567.

16. Gheblawi M, Wang K, Viveiros A, et al. Angiotensin converting enzyme 2: SARS-CoV-2 receptor and regulator of the Renin-Angiotensin System. Circ Res 2020; Online ahead of print.

17. He FJ, MacGregor GA. Salt reduction lowers cardiovascular risk: meta-analysis of outcome trials. Lancet 2011; 378: 380–82.

18. Mozaffarian D, Fahimi S, Singh GM, et al. Global sodium consumption and death from cardiovascular causes. N Engl J Med 2014; 371: 624–34.

19. Majid DSA, Prieto MC, Navar LG. Salt-sensitive hypertension: perspectives on intrarenal mechanisms. Current Hypertension Reviews 2015; 11: 38–48.

20. Berger RC, Vassallo PF, Crajoinas Rde O, et al. Renal effects and underlying molecular mechanisms of long-term salt content diets in spontaneously hypertensive rats. PLoS One 2015; 10: e0141288.

21. Tikellis C, Pickering RJ, Tsorotes D, et al. Activation of the Renin-Angiotensin system mediates the effects of dietary salt intake on atherogenesis in the apolipoprotein E knockout mouse. Hypertension 2012; 60: 98–105.

22. Hutton B, Salanti G, Caldwell DM, et al. The PRISMA extension statement for reporting of systematic reviews incorporating network meta-analyses of health care interventions. Checklist and explanations. Ann Intern Med 2015; 162: 777–84.

23. Lin L, Li TS. Interpretation of guidelines for the diagnosis and treatment of novel coronavirus (2019-nCoV) infection by the national health commission (Trial Version 5). Zhonghua Yi Xue Za Zhi 2020; 100: E001.

24. Wrotek A, Jackowska T, Pawlik K. Sodium and copeptin levels in children with community acquired pneumonia. Adv Exp Med Biol 2015; 835: 31–6.

25. Giannis D, Matenoglou E, Moris D. Hyponatremia as a marker of complicated appendicitis: A systematic review. Surgeon 2020; S1479–666X (20)30013–5.

26. Lindestam U, Almström M, Jacks J, et al. Low plasma sodium concentration predicts perforated acute appendicitis in children: A prospective diagnostic accuracy study. Eur J Pediatr Surg 2019; Epub ahead of print.

27. WHO guideline: sodium intake for adults and children. Geneva: World Health Organization, 2012.

28. Whelton PK, Appel LJ, Sacco RL, et al. Sodium, blood pressure, and cardiovascular disease: further evidence supporting the American Heart Association sodium reduction recommendations. Circulation 2012;126: 2880–89.

29. Cryan PM, Meteyer CU, Blehert DS, et al. Electrolyte depletion in white-nose syndrome bats. J Wildl Dis 2013; 49: 398–02.

30. Warnecke L, Turner JM, Bollinger TK, et al. Pathophysiology of white-nose syndrome in bats: a mechanistic model linking wing damage to mortality. Biol Lett 2013; 9: 20130177.

## References for systematic review and meta-analysis

1. Wei-jie Guan, Zheng-yi Ni, Yu Hu, Wen-hua Liang, Chun-quan Ou, Jian-xing He, Lei Liu, Hong Shan, Chun-liang Lei, David SC Hui, Bin Du, Lan-juan Li, Guang Zeng, Kowk-Yung Yuen, Ru-chong Chen, Chun-li Tang, Tao Wang, Ping-yan Chen, Jie Xiang, Shi-yue Li, Jin-lin Wang, Zi-jing Liang, Yi-xiang Peng, Li Wei, Yong Liu, Ya-hua Hu, Peng Peng, Jian-ming Wang, Ji-yang Liu, Zhong Chen, Gang Li, Zhi-jian Zheng, Shao-qin Qiu, Jie Luo, Chang-jiang Ye, Shao-yong Zhu, Nan-s han Zhong. Clinical characteristics of 2019 novel coronavirus infection in China. medRxiv2020.02.06.20020974; doi: https://doi.org/10.1101/2020.02.06.20020974

2. Zhen Li, Ming Wu, Jiwei Yao, Jie Guo, Xiang Liao, Siji Song, Jiali Li, Guangjie Duan, Yuanxiu Zhou, Xiaojun Wu, Zhan song Zhou, Taojiao Wang, Ming Hu, Xianxiang Chen, Yu Fu, Chong Lei, Hailong Dong, Chuou Xu, Yahua Hu, Min Han, Yi Zhou, Hongbo Jia, Xiaowei Chen, Junan Yan. Caution on Kidney Dysfunctions of COVID-19 Patients Anti-2019-nCoV Volunteers. medRxiv2020.02.08.20021212; doi: https://doi.org/10.1101/2020.02.08.20021212

3. Jingyuan Liu, Yao Liu, Pan Xiang, Lin Pu, Haofeng Xiong, Chuansheng Li, Ming Zhang, Jianbo Tan, Yanli Xu, Rui Song, Meihua Song, Lin Wang, Wei Zhang, Bing Han, Li Yang, Xiaojing Wang, Guiqin Zhou, Ting Zhang, Ben Li, Yanbin Wang, Zhihai Chen, Xianbo Wang. Neutrophil-to-Lymphocyte Ratio Predicts Severe Illness Patients with 2019 Novel Coronavirus in the Early Stage. medRxiv2020.02.10.20021584; doi: https://doi.org/10.1101/2020.02.10.20021584

4. Jing Liu, Sumeng Li, Jia Liu, Boyun Liang, Xiaobei Wang, Hua Wang, Wei Li, Qiaoxia Tong, Jianhua Yi, Lei Zhao, Lijuan Xiong, Chunxia Guo, Jin Tian, Jinzhuo Luo, Jinghong Yao, Ran Pang, Hui Shen, Cheng Peng, Ting Liu, Qian Zhang, Jun Wu, Ling Xu, Sihong Lu, Baoju Wang, Zhihong Weng, Chunrong Han, Huabing Zhu, Ruxia Zhou, Helong Zhou, Xiliu Chen, Pian Ye, Bin Zhu, Shengsong He, Yongwen He, Shenghua Jie, Ping Wei, Jianao Zhang, Yinping Lu, Weixian Wang, Li Zhang, Ling Li, Fengqin Zhou, Jun Wang, Ulf Dittmer, Mengji Lu, Yu Hu, Dongliang Yang, Xin Zheng. Longitudinal characteristics of lymphocyte responses and cytokine profiles in the peripheral blood of SARS-CoV-2 infected patients. medRxiv2020.02.16.20023671; doi: https://doi.org/10.1101/2020.02.16.20023671

5. Yichun Cheng, Ran Luo, Kun Wang, Meng Zhang, Zhixiang Wang, Lei Dong, Junhua Li, Ying Yao, Shuwang Ge, Gang Xu. Kidney impairment is associated with in-hospital death of COVID-19 patients. medRxiv2020.02.18.20023242; doi: https://doi.org/10.1101/2020.02.18.20023242

6. Yishan Zheng, Zhen Huang, Guoping Ying, Xia Zhang, Wei Ye, Zhiliang Hu, Chunmei Hu, Hongxia Wei, Yi Zeng, Yun C hi, Cong Cheng, Feishen Lin, Hu Lu, Lingyan Xiao, Yan Song, Chunming Wang, Yongxiang Yi, Lei Dong. Study of the lymphocyte change between COVID-19 and non-COVID-19 pneumonia cases suggesting other factors besides uncontrolled inflammation contributed to multi-organ injury. medRxiv2020.02.19.20024885; doi: https://doi.org/10.1101/2020.02.19.20024885

7. Hongzhou Lu, Jingwen Ai, Yinzhong Shen, Yang Li, Tao Li, Xian Zhou, Haocheng Zhang, Qiran Zhang, Yun Ling, Sheng Wang, Hongping Qu, Yuan Gao, Yingchuan Li, Kanglong Yu, Duming Zhu, Hecheng Zhu, Rui Tian, Mei Zeng, Qiang Li, Yuanlin Song, Xiangyang Li, Jinfu Xu, Jie Xu, Enqiang Mao, Bijie Hu, Xin Li, Lei Zhu, Wenhong Zhang. A descriptive study of the impact of diseases control and prevention on the epidemics dynamics and clinical features of SARS-CoV-2 outbreak in Shanghai, lessons learned for metropolis epidemics prevention. medRxiv2020.02.19.20025031; doi: https://doi.org/10.1101/2020.02.19.20025031

8. Guo-Qing Qian, Nai-Bin Yang, Feng Ding, Ada Hoi, Yan Ma, Zong-Yi Wang, Yue-Fei Shen, Chun-Wei Shi, Xiang Lian, Jin-Guo Chu, Lei Chen, Zhi-Yu Wang, Da-Wei Ren, Guo-Xiang Li, Xue-Qin Chen, Hua-Jiang Shen, Xiao-Min Chen. Epidemiologic and Clinical Characteristics of 91 Hospitalized Patients with COVID-19 in Zhejiang, China: A retrospective, multi-centre case series. medRxiv2020.02.23.20026856; doi: https://doi.org/10.1101/2020.02.23.20026856

9. Pengfei Cui, Zhe Chen, Tian Wang, Jun Dai, Jinjin Zhang, Ting Ding, Jingjing Jiang, Jia Liu, Cong Zhang, Wanying Shan, Sheng Wang, Yueguang Rong, Jiang Chang, Xiaoping Miao, Xiangyi Ma, Shixuan Wang. Clinical features and sexual transmission potential of SARS-CoV-2 infected female patients: a descriptive study in Wuhan, China. medRxiv2020.02.26.20028225; doi: https://doi.org/10.1101/2020.02.26.20028225

10. Ru Liu, Xiaoyan Ming, Ou Xu, Jianli Zhou, Hui Peng, Ning Xiang, Jiaming Zhang, Hong Zhu. Association of Cardiovascular Manifestations with In-hospital Outcomes in Patients with COVID-19: A Hospital Staff Data. medRxiv2020.02.29.20029348; doi: https://doi.org/10.1101/2020.02.29.20029348.

11. Xu Chen, Fang Zheng, Yanhua Qing, Shuizi Ding, Danhui Yang, Cheng Lei, Zhilan Yin, Xianglin Zhou, Dixuan Jiang, Qi Zuo, Jun He, Jianlei Lv, Ping Chen, Yan Chen, Hong Peng, Honghui Li, Yuanlin Xie, Jiyang Liu, Zhiguo Zhou, Hong Luo. Epidemiological and clinical features of 291 cases with coronavirus disease 2019 in areas adjacent to Hubei, China: a double-center observational study. medRxiv2020.03.03.20030353; doi: https://doi.org/10.1101/2020.03.03.20030353

12. Yonghao Xu, Zhiheng Xu, Xuesong Liu, Lihua Cai, Haichong Zheng, Yongbo Huang, Lixin Zhou, Linxi Huang, Yun Lin, Liehua Deng, Jianwei Li, Sibei Chen, Dongdong Liu, Zhimin Lin, Liang Zhou, Weiqun He, Xiaoqing Liu, Yimin Li. Clinical findings in critical ill patients infected with SARS-Cov-2 in Guangdong Province, China: a multi-center, retrospective, observational study. medRxiv2020.03.03.20030668; doi: https://doi.org/10.1101/2020.03.03.20030668

13. Chengfeng Qiu, Qian Xiao, Xin Liao, Ziwei Deng, Huiwen Liu, Yuanlu Shu, Dinghui Zhou, Ye Deng, Hongqiang Wang, Xiang Zhao, Jianliang Zhou, Jin Wang, Zhihua Shi, Long Da. Transmission and clinical characteristics of coronavirus disease 2019 in 104 outside-Wuhan patients, China. medRxiv2020.03.04.20026005; doi: https://doi.org/10.1101/2020.03.04.20026005

14. Min Cao, Dandan Zhang, Youhua Wang, Yunfei Lu, Xiangdong Zhu, Ying Li, Honghao Xue, Yunxiao Lin, Min Zhang, Yiguo Sun, Zongguo Yang, Jia Shi, Yi Wang, Chang Zhou, Yidan Dong, Ping Liu, Steven M Dudek, Zhen Xiao, Hongzhou Lu, Longping Peng. Clinical Features of Patients Infected with the 2019 Novel Coronavirus (COVID-19) in Shanghai, China. medRxiv2020.03.04.20030395; doi: https://doi.org/10.1101/2020.03.04.20030395

15. Xu-wei Hong, Ze-pai Chi, Guo-yuan Liu, Hong Huang, Shun-qi Guo, Jing-ru Fan, Xian-wei Lin, Liao-zhun Qu, Rui-lie Chen, Ling-jie Wu, Liang-yu Wang, Qi-chuan Zhang, Su-wu Wu, Ze-qun Pan, Hao Lin, Yu-hua Zhou, Yong-hai Zhang. Analysis of early renal injury in COVID-19 and diagnostic value of multi-index combined detection. medRxiv2020.03.07.20032599; doi: https://doi.org/10.1101/2020.03.07.20032599

16. Yongyu Liu, Lijun Bi, Yu Chen, Yaguo Wang, Joy Fleming, Yanhong Yu, Ye Gu, Chang Liu, Lichao Fan, Xiaodan Wang, Moxin Cheng. Active or latent tuberculosis increases susceptibility to COVID-19 and disease severity. medRxiv2020.03.10.20033795; doi: https://doi.org/10.1101/2020.03.10.20033795

17. Che Zhang, Jiaowei Gu, Quanjing Chen, Na Deng, Jingfeng Li, Li Huang, Xihui Zhou. Clinical Characteristics of 34 Children with Coronavirus Disease-2019 in the West of China: a Multiple-center Case Series. medRxiv2020.03.12.20034686; doi: https://doi.org/10.1101/2020.03.12.20034686

18. Ke Hu, Yang Zhao, Mengmei Wang, Qiqi Zeng, Xiaorui Wang, Ming Wang, Zhishui Zheng, Xiaochen Li, Yunting Zhang, Tao Wang, Shaolin Zeng, Yan Jiang, Dan Liu, Wenzhen Yu, Fang Hu, Hongyu Qin, Jingcan Hao, Jian Yuan, Rui Shang, Meng Jiang, Xi Ding, Binghong Zhang, Bingyin Shi, Chengsheng Zhang. Identification of a super-spreading chain of transmission associated with COVID-19. medRxiv2020.03.19.20026245; doi: https://doi.org/10.1101/2020.03.19.20026245

19. Xiaomin Luo, Hongxia Xia, Weize Yang, Benchao Wang, Tangxi Guo, Jun Xiong, Zongping Jiang, Yu Liu, Xiaojie Yan, Wei Zhou, Lu Ye, Bicheng Zhang. Characteristics of patients with COVID-19 during epidemic ongoing outbreak in Wuhan, China. medRxiv2020.03.19.20033175; doi: https://doi.org/10.1101/2020.03.19.20033175

20. Shijiao Yan, Xingyue Song, Feng Lin, Haiyan Zhu, Xiaozhi Wang, Min Li, Jianwen Ruan, Changfeng Lin, Xiaoran Liu, Qiang Wu, Zhiqian Luo, Wenning Fu, Song Chen, Yong Yuan, Shengxing Liu, Jinjian Yao, Chuanzhu Lv. Clinical Characteristics of Coronavirus Disease 2019 in Hainan, China. medRxiv2020.03.19.20038539; doi: https://doi.org/10.1101/2020.03.19.20038539

21. Congliang Miao, Jinqiang Zhuang, Mengdi Jin, Huanwen Xiong, Peng Huang, Qi Zhao, Li Miao,Jiang Du, Xinying Yang, Peijie Huang, Jiang Hong. A comparative multi-centre study on the clinical and imaging features of comfirmed and uncomfirmed patients with COVID-19. medRxiv2020.03.22.20040782; doi: https://doi.org/10.1101/2020.03.22.20040782

22. Kaiyan Li, Dian Chen, Shengchong Chen, Yuchen Feng, Chenli Chang, Zi Wang, Nan Wang, Guohua Zhen. Radiographic Findings and other Predictors in Adults with Covid-19. medRxiv2020.03.23.20041673; doi: https://doi.org/10.1101/2020.03.23.20041673

23. Samuel J. S. Rubin, Samuel Robert Falkson, Nicholas Degner, Catherine Blish. Clinical characteristics associated with COVID-19 severity in California. medRxiv2020.03.27.20043661; doi: https://doi.org/10.1101/2020.03.27.20043661

24. Naibin Yang, Yuefei Shen, Chunwei Shi, Ada Hoi, Yan Ma, Xie Zhang, Xiaomin Jian, Liping Wang, Jiejun Shi, Chunyang Wu, Guoxiang Li, Yuan Fu, Keyin Wang, Mingqin Lu, Guoqing Qian. In-flight Transmission Cluster of COVID-19: A Retrospective Case Series. medRxiv2020.03.28.20040097; doi: https://doi.org/10.1101/2020.03.28.20040097

25. Suyan Tian, Xuetong Zhu, Xuejuan Sun, Jinmei Wang, Qi Zhou, Chi Wang, Li Chen, Jiancheng Xu. Longitudinal analysis of laboratory findings during the process of recovery for patients with COVID-19. medRxiv2020.04.04.20053280; doi: https://doi.org/10.1101/2020.04.04.20053280

26. Zhichao Feng, Jennifer Li, Shanhu Yao, Qizhi Yu, Wenming Zhou, Xiaowen Mao, Huiling Li, Wendi Kang, Xin Ouyang, Ji Mei, Qiuhua Zeng, Jincai Liu, Xiaoqian Ma, Pengfei Rong, Wei Wang. The Use of Adjuvant Therapy in Preventing Progression to Severe Pneumonia in Patients with Coronavirus Disease 2019: A Multicenter Data Analysis. medRxiv2020.04.08.20057539; doi: https://doi.org/10.1101/2020.04.08.20057539

27. Jin-Kui Yang, Jian-Min Jin, Shi Liu, Peng Bai, Wei He, Fei Wu, Xiao-Fang Liu, De-Min Han. Blood glucose is a representative of the clustered indicators of multi-organ injury for predicting mortality of COVID-19 in Wuhan, China. medRxiv2020.04.08.20058040; doi: https://doi.org/10.1101/2020.04.08.20058040

28. changzheng wang, Chengbin Li. Preliminary study to identify severe from moderate cases of COVID-19 using NLR&RDW-SD combination parameter. medRxiv2020.04.09.20058594; doi: https://doi.org/10.1101/2020.04.09.20058594

29. Xiufeng Jiang, Jianxin Tao, Hui Wu, Yixin Wang, Wei Zhao, Min Zhou, Jiehui Huang, Qian You, Hua Meng, Feng Zhu, Xiaoqing Zhang, Meifang Qian, Yuanwang Qiu. Clinical features and management of severe COVID-19: A retrospective study in Wuxi, Jiangsu Province, China. medRxiv2020.04.10.20060335; doi: https://doi.org/10.1101/2020.04.10.20060335

30. Xingdong Chen, Zhenqiu Liu. Early prediction of mortality risk among severe COVID-19 patients using machine learning. medRxiv2020.04.13.20064329; doi: https://doi.org/10.1101/2020.04.13.20064329

31. Ishan Paranjpe, Adam Russak, Jessica K De Freitas, Anuradha Lala, Riccardo Miotto, Akhil Vaid, Kipp W Johnson, Matteo Danieletto, Eddye Golden, Dara Meyer, Manbir Singh, Sulaiman Somani, Sayan Manna, Udit Nangia, Arjun Kapoor, Ross O’Hagan, Paul F O’Reilly, Laura M Huckins, Patricia Glowe, Arash Kia, Prem Timsina, Robert M Freeman, Matthew A Levin, Jeffrey Jhang, Adolfo Firpo, Patricia Kovatch, Joseph Finkelstein, Judith A Aberg, Emilia Bagiella, Carol RHorowitz, Barbara Murphy, Zahi A Fayad, Jagat Narula, Eric J Nestler, Valentin Fuster, Carlos Cordon-Cardo, Dennis S Charney, David L Reich, Allan C Just, Erwin P Bottinger, Alexander WCharney, Benjamin S Glicksberg, Girish Nadkarni, Mount Sinai Covid Informatics Center (MSCIC). Clinical Characteristics of Hospitalized Covid-19 Patients in New York City medRxiv2020.04.19.20062117; doi: https://doi.org/10.1101/2020.04.19.20062117

32. Puyu Shi, Guoxia Ren, Jun Yang, Zhiqiang Li, Shujiao Deng, Miao Li, Shasha Wang, Xiaofeng Xu, Fuping Chen, Yuanjun Li, Chunyan Li, Xiaohua Yang, Zhaofeng Xie, Zhengxia Wu, Mingwei Chen. Clinical characteristics of imported and second-generation COVID-19 cases outside Wuhan, China: A multicenter retrospective study. medRxiv2020.04.19.20071472; doi: https://doi.org/10.1101/2020.04.19.20071472

33. Mohammad Ali Ashraf, Nasim Shokouhi, Elham Shirali, Fateme Davari-tanha, Omeed Memar, Alireza Kamalipour, Ayein Azarnoush, Avin Mabadi, Adele Ossareh, Milad Sanginabadi, Talat Mokhtari Azad, Leila Aghaghazvini, Sara Ghaderkhani, Tahereh Poordast, Alieh Pourdast, Pershang Nazemi. COVID-19 in Iran, a comprehensive investigation from exposure to treatment outcomes. medRxiv2020.04.20.20072421; doi: https://doi.org/10.1101/2020.04.20.20072421

34. Anuradha Lala, Kipp W Johnson, Adam J Russak, Ishan Paranjpe, Shan Zhao, Sulaiman Solani, Akhil Vaid, Fayzan Chaudhry, Jessica K De Freitas, Zahi A Fayad, Sean P Pinney, Matthew Levin, Alexander Charney, Emilia Bagiella, Jagat Narula, Benjamin S Glicksberg, Girish Nadkarni, James Januzzi, Donna M Mancini, Valentin Fuster. Prevalence and Impact of Myocardial Injury in Patients Hospitalized with COVID-19 Infection. medRxiv2020.04.20.20072702; doi: https://doi.org/10.1101/2020.04.20.20072702

35. Zhang X, Cai H, Hu J, Lian J, Gu J, Zhang S, Ye C, Lu Y, Jin C, Yu G, Jia H, Zhang Y, Sheng J, Li L, Yang Y. Epidemiological, clinical characteristics of cases of SARS-CoV-2 infection with abnormal imaging findings. Int J Infect Dis. 2020 Mar 20;94:81–87. doi: 10.1016/j.ijid.2020.03.040.

36. Todd J. Levy, Safiya Richardson, Kevin Coppa, Douglas P. Barnaby, Thomas McGinn, Lance B.Becker, Karina W. Davidson, Jamie S. Hirsch, Theodoros Zanos, Northwell COVID-19 Research Consortium. Estimating Survival of Hospitalized COVID-19 Patients from Admission Information. medRxiv2020.04.22.20075416; doi: https://doi.org/10.1101/2020.04.22.20075416

